# Cost effective sequencing enables longitudinal profiling of clonal hematopoiesis

**DOI:** 10.1101/2022.01.31.22270028

**Authors:** Md Mesbah Uddin, Ying Zhou, Alexander G. Bick, Bala Bharathi Burugula, Sidd Jaiswal, Pinkal Desai, Shelly-Ann Love, Kate Hayden, JoAnn Manson, Eric Whitsel, Charles Kooperberg, Pradeep Natarajan, Alexander P. Reiner, Jacob Kitzman

**Affiliations:** Cardiovascular Research Center, Massachusetts General Hospital, Boston, MA; Program in Medical and Population Genetics and the Cardiovascular Disease Initiative, Broad Institute of Harvard and MIT, Cambridge, MA; Department of Medicine, Harvard Medical School, Boston, MA; Division of Public Health Sciences, Fred Hutchinson Cancer Research Center, Seattle, WA; Division of Genetic Medicine, Department of Medicine, Vanderbilt University Medical Center, Nashville, TN; Department of Human Genetics, University of Michigan, Ann Arbor, MI; Department of Pathology, Stanford University School of Medicine, Stanford, CA; Weill Cornell Medicine, NY, USA; Department of Epidemiology, University of North Carolina, Gillings School of Global Public Health, Chapel Hill, NC; Wake Forest School of Medicine, Winston-Salem, NC, USA; Department of Medicine, Brigham and Women’s Hospital, Harvard Medical School, Boston, MA; Department of Epidemiology, University of Washington, Seattle, WA 98109, USA

## Abstract

Clonal hematopoiesis of indeterminate potential (CHIP), the age-related expansion of mutant hematopoietic stem cells, confers risk for multiple diseases of aging including hematologic cancer and cardiovascular disease. Whole-exome or genome sequencing can detect CHIP, but due to its high cost, most population studies have been cross-sectional, sequencing only a single timepoint. Here we describe a cost-effective sequencing assay for detecting CHIP. We validate this technology on a set of 548 longitudinal and multi-timepoint samples from 182 participants in the Women’s Health Initiative cohort over median 16 years, including 85 participants with ≥3 timepoints assayed. The majority (52.1%) of clonal mutations expanded over time (with a median doubling period of 7.43 years), with the others remaining static or decreasing in size in the absence of any cytotoxic therapy. This assay provides a cost-effective and sensitive platform for investigating the associations between CHIP dynamics and health outcomes at a biobank scale.

## INTRODUCTION

Chronological age is the dominant risk factor for cancers and cardiovascular disease – the leading causes of death worldwide (*1*). Aging is also associated with a higher prevalence of acquired somatic mutations, especially in frequently regenerating cells, such as hematopoietic stem cells (HSC). Clonal hematopoiesis of indeterminate potential (CHIP) is the age-related expansion (defined as variant allele fraction, VAF >2%) of cancer-associated somatic mutations (typically in *DNMT3A, TET2, ASXL1, JAK2*) in hematopoietic stem cells in the absence of unexplained cytopenia, dysplasia, or neoplasia (*2*). Recent whole exome sequence (WES) and whole genome sequence (WGS) analyses of blood-derived DNA have shown that CHIP is increasingly common with advancing age (i.e., approximately 10% of asymptomatic adults older than 70 years of age) (*3–6*). While CHIP is a risk factor for hematologic malignancy and all-cause mortality (*3, 7, 8*), a number of analyses have shown an association with atherosclerotic cardiovascular disease (*4, 9, 10*). CHIP is also associated with heightened risk of therapy-related myeloid malignancies (*11–14*). These studies underline the importance of CHIP as a novel biomarker for early detection and monitoring of multiple age-related diseases (*15, 16*). However, further longitudinal studies are needed for a better understanding of the root causes of CHIP, surveillance strategies, and how CHIP dynamics influence the development of chronic diseases.

Sensitivity for the detection of driver mutations is highly dependent on sequencing depth. Both WGS and WES are suitable for the detection of larger clones (e.g., VAF >5% in WGS (*6, 7*), and VAF >3% in WES (*3, 4*)). By comparison, deeper coverage, error-corrected targeted sequencing techniques are capable of detecting very small clones (*8, 15*), which are nearly ubiquitous in healthy adults (*17*). Additional studies of apparently healthy adults characterizing longitudinal changes in clone size over time may reveal genetic and environmental factors promoting clonal stability versus progression and yield new insights into mechanisms underlying somatic mutagenesis and aging as well as resultant disease pathogenesis and disease prediction.

Here we present a single-molecule molecular inversion probe sequencing (smMIPS) assay (*18*), that leverages a cost-effective, ultrasensitive, high-throughput targeted sequencing technique, for the detection of CHIP mutations. We apply this assay to a set of longitudinal peripheral blood DNA samples obtained over a median range of 16 years from 182 post-menopausal women from the Women’s Health Initiative to compare to whole genome sequence analysis and evaluate clonal dynamics.

## RESULTS

### CHIP panel design and validation

We designed a smMIPS capture panel tiling all coding exons (±5 bp) across the 11 most common CHIP genes, along with mutational hotspots in four other genes (Fig. 1 and Table 1). The final capture included 3526 probes, each containing a 9-mer unique molecular index (UMI) for duplicate read removal, spanning a total of 35.2 kb of genomic sequence in the target region. To validate this panel, we first re-sequenced five HapMap lymphoblastoid cell lines (LCLs) and successfully identified all variants defined by 1000G WGS datasets in the target region (n=152), with no additional variants called. Focusing on positions invariant in these cell lines, we estimate a low sequencing error rate of 0.045% (~1/2200 bp; Supplementary Note). Next, to mimic CHIP mutations across a range of variant allele fractions (VAF), we mixed these cell lines’ genomic DNAs at known proportions, and sequenced the mixture. We detected all variants present in this mixture, at allelic fractions tightly correlated with those expected given the cell lines’ mixing proportions (Pearson’s r=0.998; Fig. 2).

**Table 1.** Genes targeted on smMIPs assay and proportion of CHIP in population covered by these target regions.

| Gene name | Target size (bp) | <u>MIPS read depth</u> |  | % of CHIP in Population<sup>+</sup> |
| --- | --- | --- | --- | --- |
|  |  | Median | Mean |  |
| *ASXL1* | 4687 | 2054 | 3891 | 7.41% |
| *CBL* | 2803 | 4880 | 6845 | 0.62% |
| *DNMT3A* | 3071 | 3163 | 4325 | 45.97% |
| *GNB1* | 1023 | 4671 | 6139 | 0.97% |
| *PPM1D* | 1894 | 3889 | 4856 | 4.01% |
| *SF3B1* | 4045 | 3936 | 5073 | 2.46% |
| *TET2* | 6165 | 2033 | 3587 | 19.13% |
| *TP53* | 1714 | 4063 | 5543 | 1.97% |
| *U2AF1* | 880 | 4403 | 6126 | 0.23% |
| *ZBTB33* | 2019 | 1983 | 2632 | 2.22% |
| *ZNF318* | 6918 | 3240 | 4765 | 1.80% |
| *SRSF2* * | 1 | 289 | 512 | 1.88% |
| *IDH1* * | 1 | 1325 | 1797 | 0.09% |
| *IDH2* * | 1 | 3029 | 3909 | 0.30% |
| *JAK2* * | 1 | 3839 | 4944 | 1.65% |
| **Overall** | **35223 bp** | **2803** | **4480** | 88.83% |

\**SRSF2*, *IDH1*, *IDH2*, *JAK2* target a single hotspot mutation
+Population frequencies of CHIP previously reported in NHLBI TOPMed cohorts (6)

**Fig. 1.**
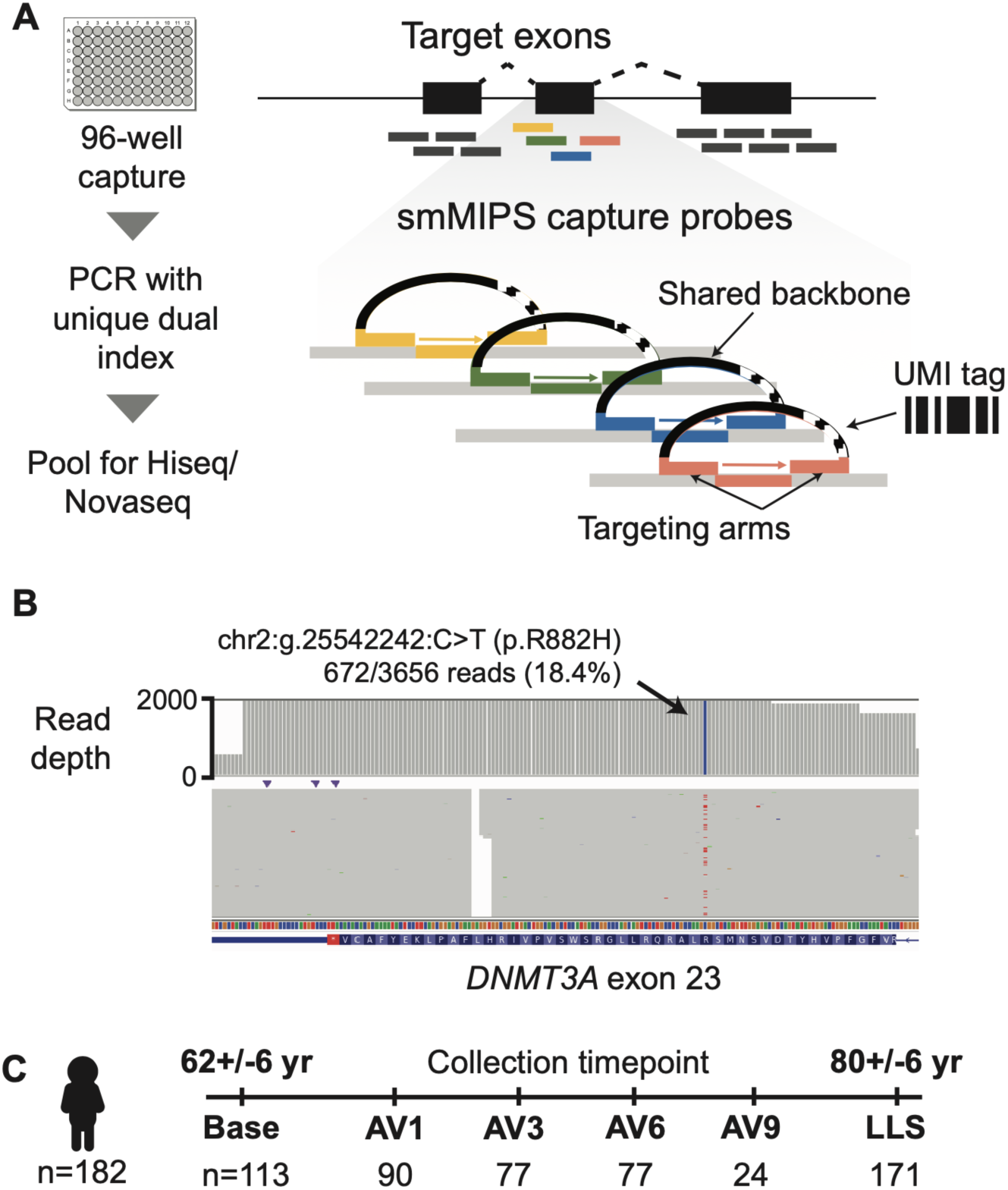
Study design and CHIP sequencing strategy. (**A**) Schematic of smMIPS assay design.(**B**) Somatic mutation identified as CHIP by smMIPS assay. (**C**) Schematic of study design, with sequencing of each subject (n=182) using samples collected at up to six timepoints including a baseline visit, a series of annual visits (AV), and a final visit (LLS, Long Life Study).

**Fig. 2.**
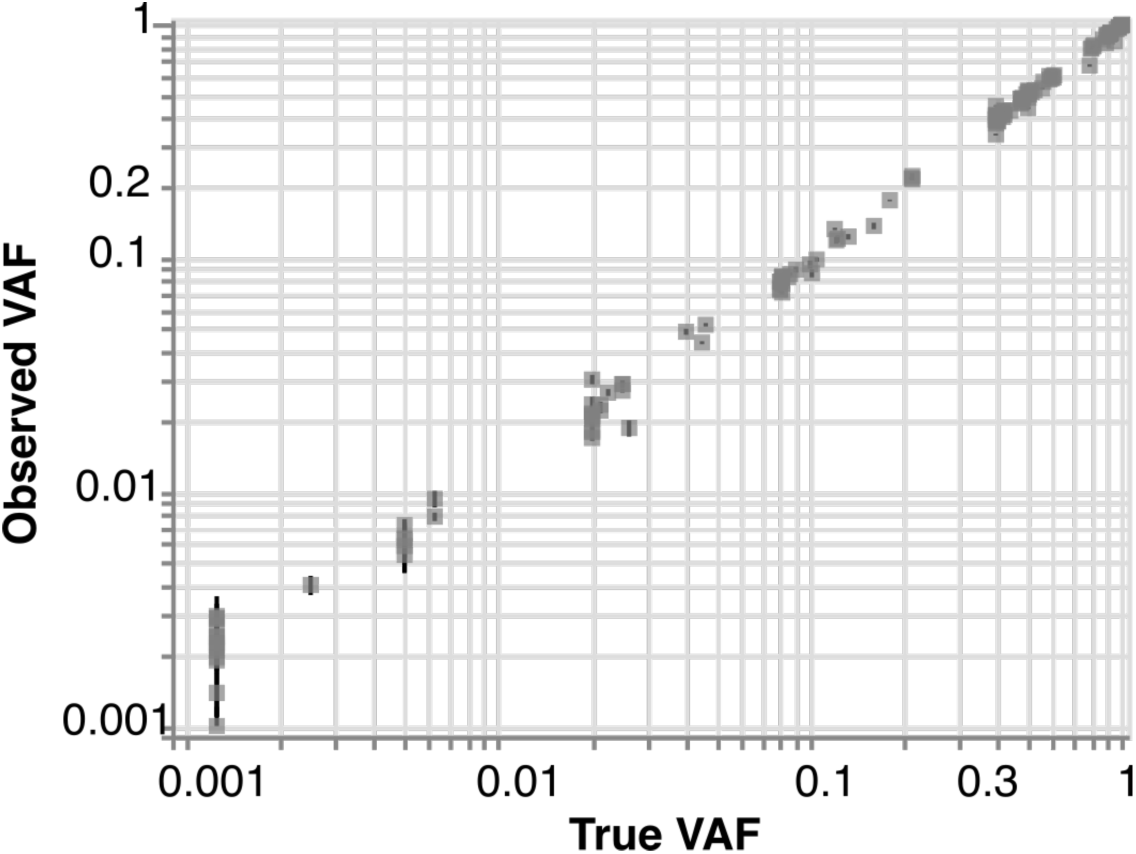
Validation by sequencing defined sample mixtures. Observed (mean +/−s.e. across 27 replicates) vs expected variant allele frequency (VAF) for repeated smMIPS sequencing of a defined control mixture of gDNAs from five cell lines, across 152 polymorphic sites. Overall Pearson’s correlation r=0.998, and r=0.847 and r=0.997 for variants with expected VAF ≤2 and >2%, respectively.

### CHIP prevalence in WHI Samples

We applied this CHIP sequencing panel to samples collected longitudinally from 182 subjects in the Women’s Health Initiative (mean: 3.0, range: 1-6 samples per subject; summarized in table S1). We obtained an overall median sequencing depth of 2803 (Table 1). After filtering, we detected a total of 178 CHIP mutations (defined as VAF ≥2%; Table S2), a subset of which were corroborated by previous whole-genome or exome sequencing of the same individuals (fig. S1). Overall, at the first timepoint sampled for each individual, 69/182 (38%) carried at least one CHIP mutation and 27/182 (15%) carried two or more mutations (Fig. 3A).

**Fig. 3.**
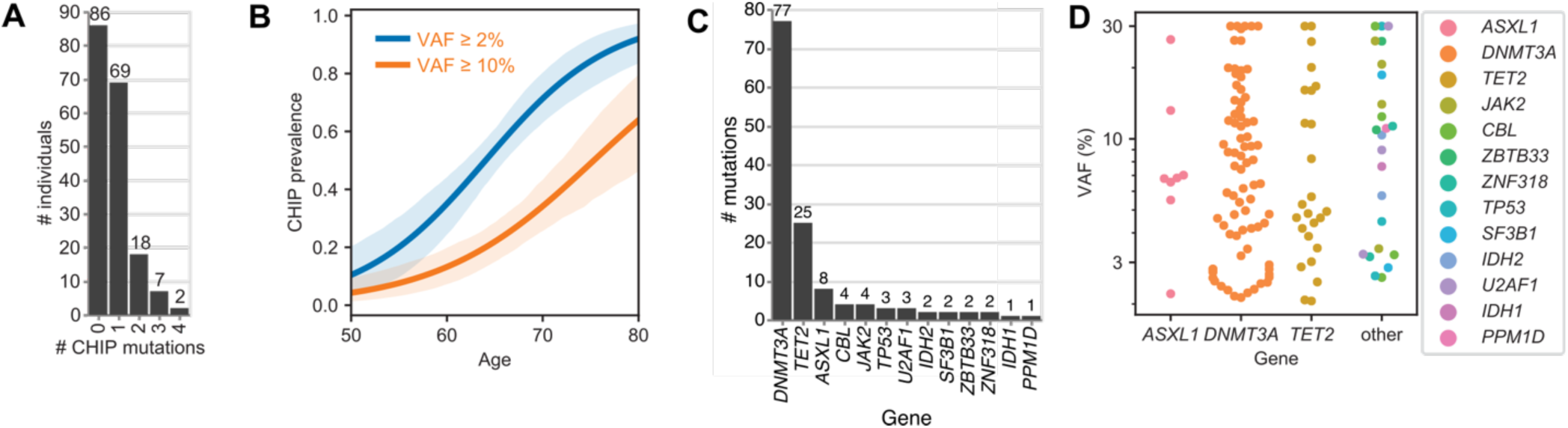
CHIP at initial blood draw. (A) Number of CHIP clones (VAF≥2%) identified per subject at initial draw. (B) Prevalence of CHIP at VAF ≥2% (blue) or ≥10% (orange) by age at initial draw. (C) Number of mutations (VAF≥2%) per gene. (D) CHIP VAFs grouped by gene.

As expected, the prevalence of CHIP (VAF ≥2%) increased with age (Fig. 3B), from 18% in individuals with initial samples taken at age 60 years or younger, compared to 84% among individuals with initial samples taken at 70 years or older, with the high prevalence reflecting in part the selection criteria for subjects previously known to be CHIP-positive at baseline. We observed BMI is significantly associated with age-adjusted CHIP VAF (p-value=0.0446), and no association with other available participant characteristics (race/ethnicity, smoking status) in this sample (Table S3).

At the baseline timepoint, the most frequently mutated genes were *DNMT3A* (57% of CHIP mutations), *TET2* (19%), and *ASXL1* (6%), consistent with prior WES or WGS reports (*4, 6*) (Fig. 3C). Among these were recurrent mutations at known hotspots including *DNMT3A* R882H/R882C (n=8 individuals) and *JAK2* V617F (n=10 individuals). In aggregate, clone sizes estimated by VAF were not significantly different by the gene mutated (Fig. 3D).

### CHIP dynamics in longitudinal samples

Of the 85 individuals with >1 draws for which CHIP (VAF≥2%) was not detected in the initial sample, 49 (58%) developed at least one CHIP mutation (VAF≥2%) at the final sampling point an average of 13.9 years later. While these late-arising clones tended to remain small (only 11/49 reached VAF≥10%), many were detectable above background at earlier time points even though they did not meet the working definition of CHIP. We classified the trajectories of each CHIP mutation, focusing on individuals (n=65) for which there were three or more timepoints, with a VAF≥1% clone in at least one of them. In these individuals, we identified 146 ‘trackable’ mutations, of which 76 were on growing trajectories, 30 were shrinking, and 40 remained static (Fig. 4). Among the mutations with a growing trajectory, the median rate of growth was 7.43 years (interquartile range: 4.48, 10.9 years) per doubling.

**Fig. 4.**
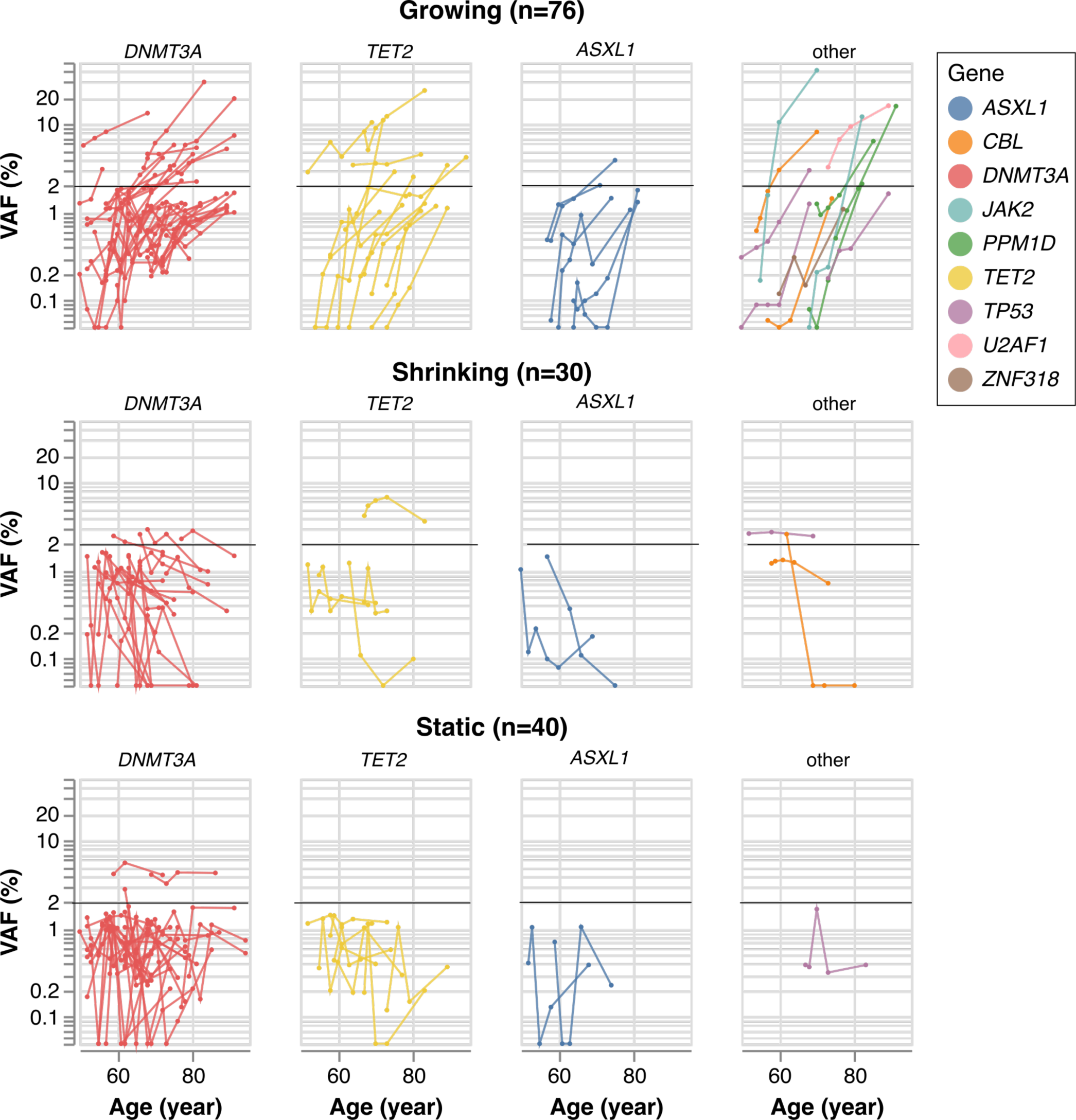
Longitudinal measurement of CHIP dynamics. Trajectories are shown, grouped by direction (rows) and gene (columns). Each trajectory corresponds to a single CHIP mutation in one subject, shaded by gene; black horizontal represents the VAF=2% threshold.

Once mutations reached appreciable frequency, they tended to continue growing, indicating that after CHIP is acquired, it is generally not lost. Among the 76 growing trajectories, 34 (44.7%) reached the VAF threshold of ≥2% and 16 of these (21.1%) reached a VAF of ≥5%. By contrast, among the shrinking or static trajectories, only 10 and 2 mutations reached these respective VAF thresholds at any single timepoint. Of the 10 non-growing clones that reached VAF≥2% at any timepoint, half involved another, growing trajectory detected in the same individual, and likely reflect competition from separate, fitter clones (Fig. 5).

**Fig. 5.**
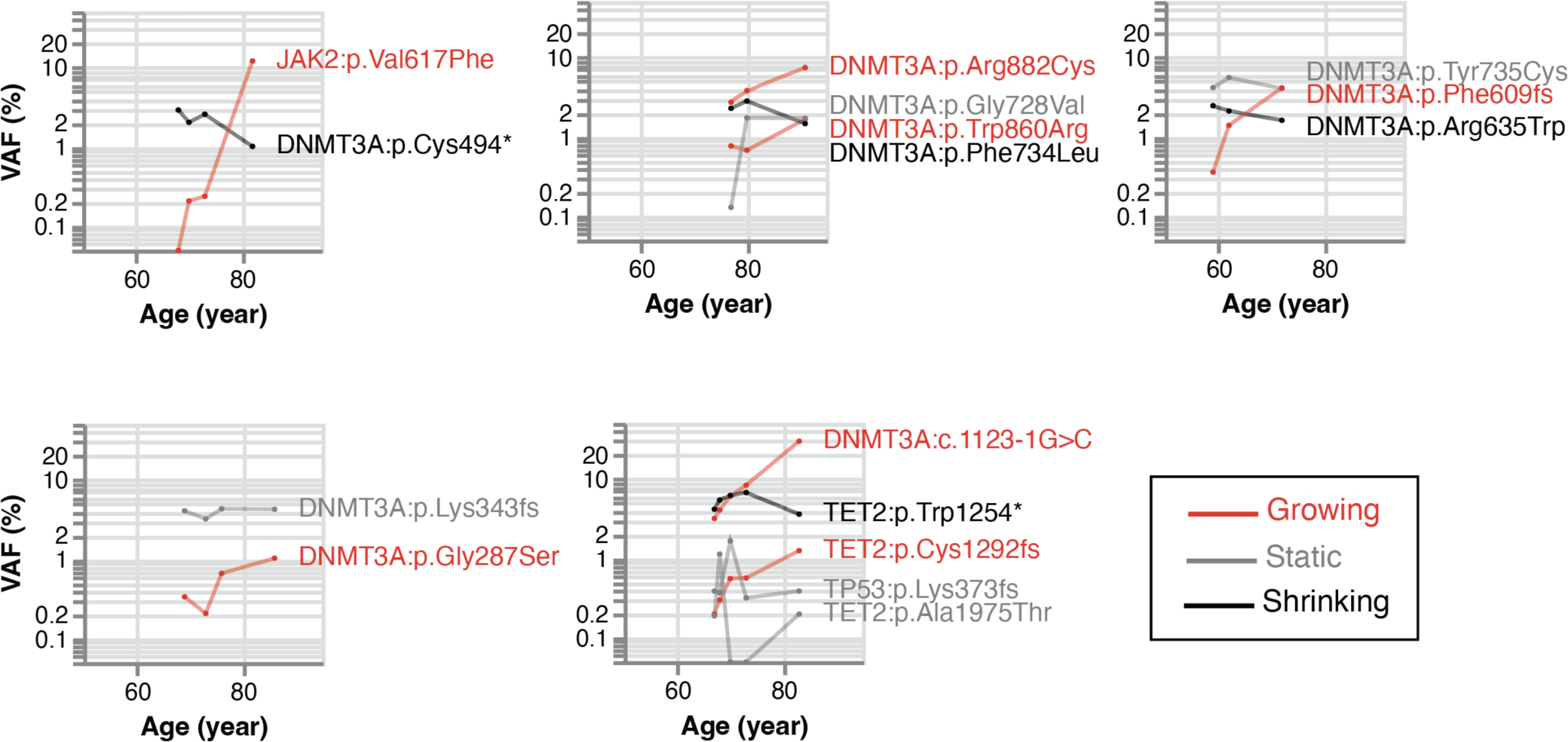
Competition between multiple different CHIP mutations. Each panel represents a single subject, and each mutation is a single line shaded by trajectory (red: growing, gray: static, black: shrinking).

### Growth rate varies by driver gene

CHIP mutations confer differential fitness advantages depending upon the gene mutated (*19–22*). To examine this, we selected the dominant (largest VAF) trajectory from each individual, removing individuals whose dominant clone trajectories are shrinking or static, leading to 43 independent trajectories from 8 CH driver genes (Table 2). Due to the small number of clones for all but the 3 major CHIP driver genes (*DNMT3A, TET2, ASXL1*; n=35 trajectories), we grouped trajectories for the other genes into a single “Other” category (n=8). The rate of growth was higher among the “other” group, which included *CBL, JAK2, TP53*, and *U2AF1*, compared to the three major CHIP genes (*P*=0.0013, Mann-Whitney U test). In addition, *DNMT3A* mutant clones were less likely to be in growing trajectories compared with other driver genes (OR=0.52, *P*=0.0085). Similar trends held among participants with only two timepoints, in which CHIP clones were classified as growing or non-growing. No association was observed between age-adjusted change in VAF (after log10 transformation) of dominant clones and any participant characteristics (race/ethnicity, smoking status and BMI) in this sample (Table S4).

**Table 2.** Number of trajectories for each driver gene.

| Gene |  | *DNMT3A* | *TET2* | *ASXL1* | *CBL* | *JAK2* | *PPM1D* | *TP53* | *U2AF1* | Total |
| --- | --- | --- | --- | --- | --- | --- | --- | --- | --- | --- |
| Growing/dominant | 23 |  | 9 | 3 | 2 | 2 | 2 | 1 | 1 | 43 |
| Growing/non-dominant |  | 20 | 6 | 3 | 0 | 0 | 1 | 2 | 0 | 33 |
| Shrinking |  | 20 | 5 | 2 | 2 | 0 | 0 | 1 | 0 | 30 |
| Static |  | 30 | 7 | 2 | 0 | 0 | 0 | 1 | 0 | 40 |

### Flexible Extension of smMIPS Platform

A key feature of the smMIPS platform is the ability to flexibly extend the probe pool to add new targets or improve coverage at existing ones. We demonstrated that here by spiking in probes at a mutational hotspot in *SRSF2*, which increased its mean coverage from <1 to 508. In addition, we added probes to target the *IL6R* p.Asp358Ala polymorphism, which stratifies the risk of cardiovascular disease for individuals with CHIP (*10*).

## DISCUSSION

Here we describe a rapid and cost-effective smMIPS-based assay that enables detection of CHIP in large scale longitudinal populations. Applying this assay to multi-time point samples from WHI participants demonstrates robust real-world performance in a large collection of longitudinal samples and reveals novel insights on clonal dynamics in a population without hematologic malignancy.

Most studies of CHIP to date rely upon either WGS or WES, or commercial capture kits which have high sequencing or library preparation costs, respectively, ranging from $150-$1000 per sample. The smMIPS approach offers a sensitive alternative at much lower per-sample cost ($30 per sample). Previous work using smMIPS for CHIP detection has been focused on individual hotspots (*23*) with full gene tiling of only *DNMT3A* (*21*). Our results demonstrate that smMIPS can scale to fully tile gene sets which cumulatively account for nearly 90% of CHIP as determined by WGS.

Our application of the smMIPS assay to WHI reveals several important insights. First, we observe a significant fraction of CHIP exists below the ~2% limit of detection afforded by conventional WES approaches to detect CHIP (*4*). The disease implications of these small clones remain to be determined in future work, enabled through cost-efficient sequencing via assays like the one described here. Second, we find that CHIP clones in the non-malignant population do not inexorably grow: just over half of those observed did expand, with the remaining, mostly low-frequency clones divided roughly evenly between static and shrinking trajectories. Future work will be required to identify the genetic and environmental factors contributing to the differing outcomes of clonal competition.

Our results add to recent observations regarding the longitudinal dynamics of clonal hematopoiesis. Fabre et al (*24*) found that clonal growth rate varies according to both age and driver gene mutation, with *DNMT3A* having a comparatively slow growth rate at older ages. In a longitudinal study of ultra-sensitive smMIP-based targeted gene sequencing of obese individuals, Van Deuren et al (*21*) reported that metabolic factors such as insulin resistance and high density lipoprotein cholesterol may accelerate expansion of CHIP clones. Using longitudinal targeted error-corrected sequence analysis in the Lothian Birth Cohorts, Robertson et al (*22*) showed that clonal growth and fitness can differ substantially by gene and gene category, with potential implications for personalized clinical management.

Our study has several limitations. First our assay robustly targets genes which account for ~90% of CHIP present in the population, so we may be misclassifying ~10% of individuals as not-having CHIP due to omission from the sequencing platform. This tradeoff was required to make the platform highly cost effective. Second, the availability of multi-time point samples is not uniform due to differences in the WHI study protocol. Third there are other kinds of clonal hematopoiesis, such as mosaic chromosomal abnormalities (e.g. structural variants) that are not detected with our CHIP assay. These limitations are balanced by the significant strengths of the novel CHIP detection assay applied to one of the largest sample sizes studied to date.

In conclusion, our development of a novel smMIPS assay for CHIP detection enables scalable and cost-effective identification of CHIP in longitudinal multi-timepoint samples from WHI. This data enabled new observations on the spectrum of clonal hematopoiesis and clonal dynamics. Future investigations using this assay at scale may enable understanding of causes of these clonal dynamic phenomena and how changes in CHIP dynamics relate to diseases of aging associated with clonal hematopoiesis.

## MATERIALS AND METHODS

### Samples

The Women’s Health Initiative (WHI) is a multicenter prospective study of risk factors for CVD, cancer, osteoporotic fractures, and other causes of morbidity and mortality among postmenopausal women (*25*). Between 1993 and 1998, women aged 50-79 years from forty WHI clinical centers throughout the United States (US) were enrolled. All WHI participants completed a baseline screening visit. A subset of participants also completed an annual visit (AV) at one, three, six, and nine years after randomization (AV1, AV3, AV6, AV9). An additional visit occurred between 2012 and 2013 (ranging from 14 to 19 years after enrollment) as part of the WHI Long Life Study (LLS), which recruited a subset of 7,875 surviving women ranging in age from 63 to 99 years at the time of LLS recruitment (*26*). The WHI-LLS cohort includes all former participants of the WHI hormone trials and all African American and Hispanic women. In order to maximize our ability to assess longitudinal CHIP trajectories over time, for the current smMIPS-based sequencing study, we included peripheral blood DNA samples of 182 WHI-LLS participants (without known prevalent hematological malignancy) of which 174 were from the WHI-LLS. Median age of participants was 62 years at baseline (range: 50-78 years) and 81 years (range: 66-95 years) at the LLS visit, respectively. Participants were selected on the basis of having undergone WGS-based CHIP determination through the NHLBI TOPMed project (N=100) and/or having DNA samples at multiple time points (N=86). From the WHI TOPMed participants, we intentionally over-sampled participants who were previously determined to have CHIP at the baseline exam (VAF>2% based on WGS) in order to directly compare intra-subject CHIP detection and VAF as determined by different assays. Genomic DNA was extracted from peripheral blood leukocytes using the 5 Prime DNA extraction kit. DNA samples were collected across multiple timepoints including at baseline, at years 1, 3, 6, 9 and at the LLS visit which is on average 15.4 years after baseline.

### Single molecule molecular inversion probe sequencing (smMIPS) assay

A smMIPS capture panel was designed to tile coding exons (+/−5 bp) of the 11 most common CHIP genes (*9*) and recurrent mutational hotspots in four others (Table 1). Probe sequences were selected as previously described (*27*), with adjustments to eliminate the need for custom sequencing primers. Briefly, probe libraries were synthesized as a 12k oligo pool by CustomArray (Bothell, WA) Inc, and subjected to bulk PCR amplification using flanking primers jklab0255_2019mipsPrep1f (GAGATCGGCGCGTTAGAAGAC) and jklab0256_2019mipsPrep1r (TGCAGGATCTAGGGCGAAGAC). PCR product was cleaned with 2.5X SPRI beads and eluted in 1X NEB cut smart buffer. To generate capture-ready probe pools, flanking adaptors were removed by BbsI-HF (#R3539L, NEB; Ipswitch, MA) digestion, overnight at 37^0^C. Digested probes were cleaned by incubating with 1x volume SPRI beads (supplemented with 5 volumes isopropanol for 20 minutes), followed by washes in 70% ethanol and elution in Tris-EDTA pH 8. Poorly captured regions were tiled with additional probes (N=112), synthesized as an oPool library by Integrated DNA Technologies (Coralville, IA) lacking flanking amplification adaptors and with 5’ phosphates. Original and make-up probes were combined into a single pool before use.

Capture reactions were assembled in a 96-well format, in 20 ul volume containing: probes (150:1 molar excess to genomic DNA targets), 1X Ampligase buffer, 1U Ampligase (Lucigen; Madison, WI), dNTPs at 0.4 uM each and 0.32 ul Hemo KlenTaq polymerase (NEB). Plates were incubated in a thermocycler at 95^0^C for 10 minutes, 95^0^C → 60^0^C at −0.1^0^C/sec, followed by a hold at 60^0^C for 18 – 24 hours. Exonuclease treatment was continued immediately after capture by adding 2 ul of mix containing 1X Ampligase buffer, 5U Exonuclease I (NEB), and 25U Exonuclease III (NEB) to each sample. Reactions were incubated at 37^0^C for 45 minutes and 95^0^C for 2 minutes. Dual indexed sequencing libraries were constructed by PCR amplification using indexing primers directed against common sequences on the probe backbone. Libraries were pooled at equal volumes, purified by 0.9X SPRI beads, and sequenced in batches of 196 on Hiseq 4000 or Novaseq instruments with paired-end 150-bp reads. Reagent, consumable, and sequencing costs total approximately $30 USD/sample.

Sequencing reads were aligned to the human reference genome (build 37) with bwa mem (*28*), and a custom sequencing pipeline (https://github.com/kitzmanlab/mimips) was used for post-alignment processing to remove probe arm sequences from each alignment and filter reads with duplicate unique molecular identifiers (UMIs).

### smMIPS Assay Validation and Reliability

To validate the clone size detection limit of the smMIPS method, we prepared mixtures of gDNAs from five lymphoblastoid cell lines (GM06994, GM12878, GM20847, GM12877 and GM18507) with known genotypes, combined at 78.8%, 16%, 4%, 1%, 0.25%. Within the target region, these cell lines have 152 known variants as defined by the 1000 Genome Project (1000G) WGS genotypes and by detecting germline variants by sequencing cell lines individually. In the resulting mixture, their expected VAFs range from 0.125% to 100%. These variants constituted the true positive variant set. We also defined as ‘true negative’ sites 13 common polymorphism SNVs absent from all of the five cell lines, and those sites (+/−50bp) were defined as true negative variants. The positive control mixture was included with each sequencing batch for a total of 27 replicates. The between-variant reliability of VAF estimated as an intraclass correlation coefficient was 0.998 (95% confidence interval 0.998 to 0.999) (see Supplementary Note).

### Variant calling

Somatic SNPs and indels were called using LoFreq 2.1.3.1 (*29*), requiring minimum coverage 40, with ≥5 reads supporting the alternate allele and a variant allele frequency (VAF) ≥ 0.1%. Variants present in ≥5% of samples at a VAF of 1-10% were discarded as likely recurrent artifacts.

### CHIP calling

Variants were annotated using ANNOVAR software (*30*). Variant calls processed using an existing filtering pipeline based upon gene name, variant functional class, and populational allele frequency (*6*); workflow is available at available at https://app.terra.bio/#workspaces/terra-outreach/CHIP-Detection-Mutect2/notebooks. For *ZBTB33* and *ZNF318*, two genes not listed in (*6*), we included variants annotated as frameshift/splice-site/nonsense or nonsynonymous(*31*). The full list of specific mutations queried is presented in Table S4. We manually reviewed alignments for selected CHIP variant calls using Integrative Genomics Viewer (IGV) (*32*).

### CHIP clone trajectories

To characterize the longitudinal trajectory of each CHIP clone over time, we restricted our analysis to individuals who (a) underwent smMIPS sequencing at least 3 time points and (b) had at least one CHIP mutation detectable at VAF>1% at any of the timepoints. We excluded any variants with alternate read count <2 or total read depth <200. For each CHIP mutation meeting these criteria, we modeled the trajectory by fitting a linear regression: log10(VAF) = C + β * age; VAFs of zero were set to a minimum of 10^−4^ (reflecting a conservative limit of detection for smMIPS), and each observation was weighted by the square root of the read depth. To further characterize clonal dynamics, we classified each trajectory based on linear trajectory, as (a) growing (β>0, *P*<0.5), (b) shrinking (β≤0, *P*<0.5), or (c) static (*P*≥0.5). For trajectory analysis, we excluded CHIP clones with starting VAF >10%, for which an exponential growth assumption may not fit.

### Association between participant characteristics and CHIP VAF and growth rate

For cross-sectional analyses at a single time point, we fit linear and logistic regression models to assess the relationship of either CHIP prevalence (total clones or large clones only) or log-transformed VAF to age at blood draw, race/ethnicity, smoking status, or BMI. We used the first visit time point for each subject. To assess the relationship of these same participant characteristics, to clone growth over time, we utilized longitudinal data from all individuals with sequencing data from at least two time points with positive VAF observations (N=148 individuals). For each participant, we first selected a single driver clone, prioritizing as the predominant clone those with the highest VAF at any follow-up timepoint. We used a linear regression to determine the effect of age, race/ethnicity, smoking, or BMI on the difference (after log10 transformation) between the first non-zero VAF value and the last non-zero VAF. All statistical analyses were adjusted for the first visit time of year and performed in R version 4.2 (R Core Team, URL https://www.R-project.org/).

## Data Availability

Sequencing data in this study are deposited (in process) at dbGaP.

## Author contributions

A.G.B., A.P.R., P.N. and J.O.K. conceived and designed the study. B.B. performed DNA library preparation and sequencing. M.M.U., Y.Z., A.G.B., and J.O.K. analyzed the data. A.G.B., P.N., A.P.R., M.M.U., Y.Z., and J.O.K. wrote the manuscript. All authors received and approved the manuscript.

## Data and materials availability

Sequencing data in this study are available in the dbGaP database at accession ####### (submission in process, will be available upon publication).

## Supplementary Materials

### Supplementary Note

**Fig. S1. Comparison of VAFs for CHIP mutations detected by smMIPS (y-axis) vs VAF from WGS of the same sample (x-axis); Pearson’s r=0.79**.

**Table S1: Summary for 182 WHI subjects**.

**Table S2. CHIP calls in the WHI longitudinal samples**. AV: Annual visit; LLS: Long-life study visit; REF: Reference Allele; ALT: Alternative Allele; VAF: Variant allele fraction; ALT Count: number of reads supporting alternative allele.

**Table S3: Cross-sectional association at the first visit**. For model 1-4, logistic regressions are conducted on the prevalence of CHIP mutation (VAF >= 2% VS VAF <2%) against age and covariates race (Black/ White /Unknown or Other), smoking history (Past/ Never/ Current/ Missing), and BMI. For models 2 and 3, model 1 is the null model and p-values are obtained from a likelihood ratio test whether the covariate is associated with the prevalence of CHIP mutations. For model 5-8, linear regressions are conducted on the log10(VAF) of detectable CHIP mutation (VAF >0) against age and the covariate. For models 6 and 7, the model 5 is the null-model, and p-values are obtained from F tests.

**Table S4: Association tests of VAF changes between the first and last positive VAFs (n=148)**. Linear regressions are conducted on the log10(VAF) changes of CHIP mutation against initial age, age length (age differences between the first and the last VAF samples), and covariate. For models 3 and 4, model 2 is the null model and p-values are obtained from testing whether race/smoking is associated with the prevalence of CHIP mutations. The covariates include race categories (Black/White/Unknown or other), smoking history (Never/Ever/Missing), and average BMI. For each subject carrying multiple clones, the dominant clone is selected based on the largest VAF value at any available time points.

**Table S5**: **CHIP mutations queried in this study**. CHIP: clonal hematopoiesis of indeterminate potential; *SRSF2, IDH1, IDH2, JAK2 target a single hotspot mutation

## References

1. GBD 2016 Causes of Death Collaborators, Global, regional, and national age-sex specific mortality for 264 causes of death, 1980-2016: a systematic analysis for the Global Burden of Disease Study 2016. Lancet 390, 1151–1210 (2017).

2. D. P. Steensma, R. Bejar, S. Jaiswal, R. C. Lindsley, M. A. Sekeres, R. P. Hasserjian, B. L. Ebert, Clonal hematopoiesis of indeterminate potential and its distinction from myelodysplastic syndromes. Blood 126, 9–16 (2015).

3. G. Genovese, A. K. Kahler, R. E. Handsaker, J. Lindberg, S. A. Rose, S. F. Bakhoum, K. Chambert, E. Mick, B. M. Neale, M. Fromer, S. M. Purcell, O. Svantesson, M. Landen, M. Hoglund, S. Lehmann, S. B. Gabriel, J. L. Moran, E. S. Lander, P. F. Sullivan, P. Sklar, H. Gronberg, C. M. Hultman, S. A. McCarroll, Clonal hematopoiesis and blood-cancer risk inferred from blood DNA sequence. N Engl J Med 371, 2477–2487 (2014).

4. S. Jaiswal, P. Fontanillas, J. Flannick, A. Manning, P. V. Grauman, B. G. Mar, R. C. Lindsley, C. H. Mermel, N. Burtt, A. Chavez, J. M. Higgins, V. Moltchanov, F. C. Kuo, M. J. Kluk, B. Henderson, L. Kinnunen, H. A. Koistinen, C. Ladenvall, G. Getz, A. Correa, B. F. Banahan, S. Gabriel, S. Kathiresan, H. M. Stringham, M. I. McCarthy, M. Boehnke, J. Tuomilehto, C. Haiman, L. Groop, G. Atzmon, J. G. Wilson, D. Neuberg, D. Altshuler, B. L. Ebert, Age-related clonal hematopoiesis associated with adverse outcomes. N Engl J Med 371, 2488–2498 (2014).

5. M. Xie, C. Lu, J. Wang, M. D. McLellan, K. J. Johnson, M. C. Wendl, J. F. McMichael, H. K. Schmidt, V. Yellapantula, C. A. Miller, B. A. Ozenberger, J. S. Welch, D. C. Link, M. J. Walter, E. R. Mardis, J. F. Dipersio, F. Chen, R. K. Wilson, T. J. Ley, L. Ding, Age-related mutations associated with clonal hematopoietic expansion and malignancies. Nat Med 20, 1472–1478 (2014).

6. A. G. Bick, J. S. Weinstock, S. K. Nandakumar, C. P. Fulco, E. L. Bao, S. M. Zekavat, M. D. Szeto, X. Liao, M. J. Leventhal, J. Nasser, K. Chang, C. Laurie, B. B. Burugula, C. J. Gibson, A. E. Lin, M. A. Taub, F. Aguet, K. Ardlie, B. D. Mitchell, K. C. Barnes, A. Moscati, M. Fornage, S. Redline, B. M. Psaty, E. K. Silverman, S. T. Weiss, N. D. Palmer, R. S. Vasan, E. G. Burchard, S. L. R. Kardia, J. He, R. C. Kaplan, N. L. Smith, D. K. Arnett, D. A. Schwartz, A. Correa, M. de Andrade, X. Guo, B. A. Konkle, B. Custer, J. M. Peralta, H. Gui, D. A. Meyers, S. T. McGarvey, I. Y. Chen, M. B. Shoemaker, P. A. Peyser, J. G. Broome, S. M. Gogarten, F. F. Wang, Q. Wong, M. E. Montasser, M. Daya, E. E. Kenny, K. E. North, L. J. Launer, B. E. Cade, J. C. Bis, M. H. Cho, J. Lasky-Su, D. W. Bowden, L. A. Cupples, A. C. Y. Mak, L. C. Becker, J. A. Smith, T. N. Kelly, S. Aslibekyan, S. R. Heckbert, H. K. Tiwari, I. V. Yang, J. A. Heit, S. A. Lubitz, J. M. Johnsen, J. E. Curran, S. E. Wenzel, D. E. Weeks, D. C. Rao, D. Darbar, J. Y. Moon, R. P. Tracy, E. J. Buth, N. Rafaels, R. J. F. Loos, P. Durda, Y. Liu, L. Hou, J. Lee, P. Kachroo, B. I. Freedman, D. Levy, L. F. Bielak, J. E. Hixson, J. S. Floyd, E. A. Whitsel, P. T. Ellinor, M. R. Irvin, T. E. Fingerlin, L. M. Raffield, S. M. Armasu, M. M. Wheeler, E. C. Sabino, J. Blangero, L. K. Williams, B. D. Levy, W. H. Sheu, D. M. Roden, E. Boerwinkle, J. E. Manson, R. A. Mathias, P. Desai, K. D. Taylor, A. D. Johnson, N. T.-O. f. P. M. Consortium, P. L. Auer, C. Kooperberg, C. C. Laurie, T. W. Blackwell, A. V. Smith, H. Zhao, E. Lange, L. Lange, S. S. Rich, J. I. Rotter, J. G. Wilson, P. Scheet, J. O. Kitzman, E. S. Lander, J. M. Engreitz, B. L. Ebert, A. P. Reiner, S. Jaiswal, G. Abecasis, V. G. Sankaran, S. Kathiresan, P. Natarajan, Inherited causes of clonal haematopoiesis in 97,691 whole genomes. Nature 586, 763–768 (2020).

7. F. Zink, S. N. Stacey, G. L. Norddahl, M. L. Frigge, O. T. Magnusson, I. Jonsdottir, T. E. Thorgeirsson, A. Sigurdsson, S. A. Gudjonsson, J. Gudmundsson, J. G. Jonasson, L. Tryggvadottir, T. Jonsson, A. Helgason, A. Gylfason, P. Sulem, T. Rafnar, U. Thorsteinsdottir, D. F. Gudbjartsson, G. Masson, A. Kong, K. Stefansson, Clonal hematopoiesis, with and without candidate driver mutations, is common in the elderly. Blood 130, 742–752 (2017).

8. P. Desai, N. Mencia-Trinchant, O. Savenkov, M. S. Simon, G. Cheang, S. Lee, M. Samuel, E. K. Ritchie, M. L. Guzman, K. V. Ballman, G. J. Roboz, D. C. Hassane, Somatic mutations precede acute myeloid leukemia years before diagnosis. Nat Med 24, 1015–1023 (2018).

9. S. Jaiswal, P. Natarajan, A. J. Silver, C. J. Gibson, A. G. Bick, E. Shvartz, M. McConkey, N. Gupta, S. Gabriel, D. Ardissino, U. Baber, R. Mehran, V. Fuster, J. Danesh, P. Frossard, D. Saleheen, O. Melander, G. K. Sukhova, D. Neuberg, P. Libby, S. Kathiresan, B. L. Ebert, Clonal Hematopoiesis and Risk of Atherosclerotic Cardiovascular Disease. N Engl J Med 377, 111–121 (2017).

10. A. G. Bick, J. P. Pirruccello, G. K. Griffin, N. Gupta, S. Gabriel, D. Saleheen, P. Libby, S. Kathiresan, P. Natarajan, Genetic Interleukin 6 Signaling Deficiency Attenuates Cardiovascular Risk in Clonal Hematopoiesis. Circulation 141, 124–131 (2020).

11. N. K. Gillis, M. Ball, Q. Zhang, Z. Ma, Y. Zhao, S. J. Yoder, M. E. Balasis, T. E. Mesa, D. A. Sallman, J. E. Lancet, R. S. Komrokji, A. F. List, H. L. McLeod, M. Alsina, R. Baz, K. H. Shain, D. E. Rollison, E. Padron, Clonal haemopoiesis and therapy-related myeloid malignancies in elderly patients: a proof-of-concept, case-control study. Lancet Oncol 18, 112–121 (2017).

12. J. I. Hsu, T. Dayaram, A. Tovy, E. De Braekeleer, M. Jeong, F. Wang, J. Zhang, T. P. Heffernan, S. Gera, J. J. Kovacs, J. R. Marszalek, C. Bristow, Y. Yan, G. Garcia-Manero, H. Kantarjian, G. Vassiliou, P. A. Futreal, L. A. Donehower, K. Takahashi, M. A. Goodell, PPM1D Mutations Drive Clonal Hematopoiesis in Response to Cytotoxic Chemotherapy. Cell Stem Cell 23, 700–713 e706 (2018).

13. K. L. Bolton, R. N. Ptashkin, T. Gao, L. Braunstein, S. M. Devlin, D. Kelly, M. Patel, A. Berthon, A. Syed, M. Yabe, C. C. Coombs, N. M. Caltabellotta, M. Walsh, K. Offit, Z. Stadler, D. Mandelker, J. Schulman, A. Patel, J. Philip, E. Bernard, G. Gundem, J. E. A. Ossa, M. Levine, J. S. M. Martinez, N. Farnoud, D. Glodzik, S. Li, M. E. Robson, C. Lee, P. D. P. Pharoah, K. H. Stopsack, B. Spitzer, S. Mantha, J. Fagin, L. Boucai, C. J. Gibson, B. L. Ebert, A. L. Young, T. Druley, K. Takahashi, N. Gillis, M. Ball, E. Padron, D. M. Hyman, J. Baselga, L. Norton, S. Gardos, V. M. Klimek, H. Scher, D. Bajorin, E. Paraiso, R. Benayed, M. E. Arcila, M. Ladanyi, D. B. Solit, M. F. Berger, M. Tallman, M. Garcia-Closas, N. Chatterjee, L. A. Diaz, Jr., R. L. Levine, L. M. Morton, A. Zehir, E. Papaemmanuil, Cancer therapy shapes the fitness landscape of clonal hematopoiesis. Nat Genet 52, 1219–1226 (2020).

14. C. C. Coombs, A. Zehir, S. M. Devlin, A. Kishtagari, A. Syed, P. Jonsson, D. M. Hyman, D. B. Solit, M. E. Robson, J. Baselga, M. E. Arcila, M. Ladanyi, M. S. Tallman, R. L. Levine, M. F. Berger, Therapy-Related Clonal Hematopoiesis in Patients with Non-hematologic Cancers Is Common and Associated with Adverse Clinical Outcomes. Cell Stem Cell 21, 374–382 e374 (2017).

15. S. Abelson, G. Collord, S. W. K. Ng, O. Weissbrod, N. Mendelson Cohen, E. Niemeyer, N. Barda, P. C. Zuzarte, L. Heisler, Y. Sundaravadanam, R. Luben, S. Hayat, T. T. Wang, Z. Zhao, I. Cirlan, T. J. Pugh, D. Soave, K. Ng, C. Latimer, C. Hardy, K. Raine, D. Jones, D. Hoult, A. Britten, J. D. McPherson, M. Johansson, F. Mbabaali, J. Eagles, J. K. Miller, D. Pasternack, L. Timms, P. Krzyzanowski, P. Awadalla, R. Costa, E. Segal, S. V. Bratman, P. Beer, S. Behjati, I. Martincorena, J. C. Y. Wang, K. M. Bowles, J. R. Quiros, A. Karakatsani, C. La Vecchia, A. Trichopoulou, E. Salamanca-Fernandez, J. M. Huerta, A. Barricarte, R. C. Travis, R. Tumino, G. Masala, H. Boeing, S. Panico, R. Kaaks, A. Kramer, S. Sieri, E. Riboli, P. Vineis, M. Foll, J. McKay, S. Polidoro, N. Sala, K. T. Khaw, R. Vermeulen, P. J. Campbell, E. Papaemmanuil, M. D. Minden, A. Tanay, R. D. Balicer, N. J. Wareham, M. Gerstung, J. E. Dick, P. Brennan, G. S. Vassiliou, L. I. Shlush, Prediction of acute myeloid leukaemia risk in healthy individuals. Nature 559, 400–404 (2018).

16. K. L. Bolton, A. Zehir, R. N. Ptashkin, M. Patel, D. Gupta, R. Sidlow, E. Papaemmanuil, M. F. Berger, R. L. Levine, The Clinical Management of Clonal Hematopoiesis: Creation of a Clonal Hematopoiesis Clinic. Hematol Oncol Clin North Am 34, 357–367 (2020).

17. A. L. Young, G. A. Challen, B. M. Birmann, T. E. Druley, Clonal haematopoiesis harbouring AML-associated mutations is ubiquitous in healthy adults. Nat Commun 7, 12484 (2016).

18. J. B. Hiatt, C. C. Pritchard, S. J. Salipante, B. J. O’Roak, J. Shendure, Single molecule molecular inversion probes for targeted, high-accuracy detection of low-frequency variation. Genome Res 23, 843–854 (2013).

19. C. J. Watson, A. L. Papula, G. Y. P. Poon, W. H. Wong, A. L. Young, T. E. Druley, D. S. Fisher, J. R. Blundell, The evolutionary dynamics and fitness landscape of clonal hematopoiesis. Science 367, 1449–1454 (2020).

20. M. A. Fabre, J. G. d. Almeida, E. Fiorillo, E. Mitchell, A. Damaskou, J. Rak, V. Orrù, M. Marongiu, M. Vijayabaskar, J. Baxter, C. Hardy, F. Abascal, M. S. Chapman, N. Williams, J. Nangalia, I. Martincorena, P. J. Campbell, E. F. McKinney, F. Cucca, M. Gerstung, G. S. Vassiliou., The longitudinal dynamics and natural history of clonal haematopoiesis. bioRxiv, 2021.2008.2012.455048 (2021).

21. R. C. van Deuren, J. C. Andersson-Assarsson, F. M. Kristensson, M. Steehouwer, K. Sjöholm, P.-A. Svensson, M. Pieterse, C. Gilissen, M. Taube, P. Jacobson, R. Perkins, H. G. Brunner, M. G. Netea, M. Peltonen, B. Carlsson, A. Hoischen, L. M. S. Carlsson, Expansion of mutation-driven haematopoietic clones is associated with insulin resistance and low HDL-cholesterol in individuals with obesity. bioRxiv, 2021.2005.2012.443095 (2021).

22. N. A. Robertson, E. Latorre-Crespo, M. Terradas-Terradas, A. C. Purcell, B. J. Livesey, J. A. Marsh, L. Murphy, A. Fawkes, L. MacGillivray, M. Copland, R. E. Marioni, S. E. Harris, S. R. Cox, I. J. Deary, L. J. Schumacher, K. Kirschner, T. Chandra, Longitudinal dynamics of clonal hematopoiesis identifies gene-specific fitness effects. bioRxiv, 2021.2005.2027.446006 (2021).

23. R. Acuna-Hidalgo, H. Sengul, M. Steehouwer, M. van de Vorst, S. H. Vermeulen, L. Kiemeney, J. A. Veltman, C. Gilissen, A. Hoischen, Ultra-sensitive Sequencing Identifies High Prevalence of Clonal Hematopoiesis-Associated Mutations throughout Adult Life. Am J Hum Genet 101, 50–64 (2017).

24. M. A. Fabre, T. McKerrell, M. Zwiebel, M. S. Vijayabaskar, N. Park, P. M. Wells, R. Rad, P. Deloukas, K. Small, C. J. Steves, G. S. Vassiliou, Concordance for clonal hematopoiesis is limited in elderly twins. Blood 135, 269–273 (2020).

25. The Women’s Health Initiative Study Group, Design of the Women’s Health Initiative clinical trial and observational study. Control Clin Trials 19, 61–109 (1998).

26. The Women’s Health Initiative Study Group, Long Life Study (W64). https://sp.whi.org/studies/SitePages/Long%20Life%20Study.aspx Accessed Jan 18, 2022 (2013).

27. M. I. Perez Millan, S. A. Vishnopolska, A. Z. Daly, J. P. Bustamante, A. Seilicovich, I. Bergada, D. Braslavsky, A. C. Keselman, R. M. Lemons, A. H. Mortensen, M. A. Marti, S. A. Camper, J. O. Kitzman, Next generation sequencing panel based on single molecule molecular inversion probes for detecting genetic variants in children with hypopituitarism. Mol Genet Genomic Med, (2018).

28. H. Li, R. Durbin, Fast and accurate short read alignment with Burrows-Wheeler transform. Bioinformatics 25, 1754–1760 (2009).

29. A. Wilm, P. P. Aw, D. Bertrand, G. H. Yeo, S. H. Ong, C. H. Wong, C. C. Khor, R. Petric, M. L. Hibberd, N. Nagarajan, LoFreq: a sequence-quality aware, ultra-sensitive variant caller for uncovering cell-population heterogeneity from high-throughput sequencing datasets. Nucleic Acids Res 40, 11189–11201 (2012).

30. K. Wang, M. Li, H. Hakonarson, ANNOVAR: functional annotation of genetic variants from high-throughput sequencing data. Nucleic Acids Res 38, e164 (2010).

31. E. M. Beauchamp, M. Leventhal, E. Bernard, E. R. Hoppe, G. Todisco, M. Creignou, A. Galli, C. A. Castellano, M. McConkey, A. Tarun, W. Wong, M. Schenone, C. Stanclift, B. Tanenbaum, E. Malolepsza, B. Nilsson, A. G. Bick, J. S. Weinstock, M. Miller, A. Niroula, A. Dunford, A. Taylor-Weiner, T. Wood, A. Barbera, S. Anand, B. M. Psaty, P. Desai, M. H. Cho, A. D. Johnson, R. Loos, N. T.-O. f. P. M. Consortium, D. G. MacArthur, M. Lek, C. Exome Aggregation, D. S. Neuberg, K. Lage, S. A. Carr, E. Hellstrom-Lindberg, L. Malcovati, E. Papaemmanuil, C. Stewart, G. Getz, R. K. Bradley, S. Jaiswal, B. L. Ebert, ZBTB33 is mutated in clonal hematopoiesis and myelodysplastic syndromes and impacts RNA splicing. Blood Cancer Discov 2, 500–517 (2021).

32. J. T. Robinson, H. Thorvaldsdottir, A. M. Wenger, A. Zehir, J. P. Mesirov, Variant Review with the Integrative Genomics Viewer. Cancer Res 77, e31–e34 (2017).

