## Supplementary Note for "Cost effective sequencing enables longitudinal profiling of clonal hematopoiesis"

#### **Error rate and profile for CHIP detection by smMIPS**

To evaluate the error profile of the sequencing assay we first generated smMIPS data from a mixture of five cell lines with known genotypes. We selected ~32k positions that were expected to be devoid of germline variants in those cell lines, and empirically computed error rate for the 12 possible single-base substitutions against the background reference allele; any mismatches within these loci are assumed to be originate from damage sustained during cell culture, DNA preparation, or storage, or due to errors introduced during library preparation or sequencing. Spontaneous deamination of cytosine (C>T/G>A) was evident as elevated C>T/G>A errors in the CpG dinucleotide context. We did not observe elevated substitution rate in C>A (G>T), which is a signature of oxidative DNA damage. The overall substitution rate was 0.00045, that is, 1 error in ~2200 bp of smMIPS sequencing.

#### **Reliability of measured VAF values**

Based on the 27 positive control samples sequenced in replicate, the site-specific variance of VAF was parsed into its between-site, between-replicate and residual (technical) components using random-effects analysis of variance. The reliability of VAF was then estimated as an intra-class correlation coefficient (ICC), i.e. a ratio of the between-replicate to total variance of VAF. The variance of the trimodally distributed VAF was parsed into its between-site, within-site (between-replicate), and residual (technical) components using random-effects analysis of variance. The reliability of VAF was then estimated as a series intra-class correlation coefficients (95% confidence intervals). Between-site to total variance was 0.998 (0.998, 0.999), within-site to total variance was 0.000 (-0.216, 0.216), and residual (technical) to total variance was 0.0 02 (-0.214, 0.217). They suggested that overall, VAF was highly reliable, with nearly all of the observed variation in it attributable to that between sites.

### **Supplementary Figures**

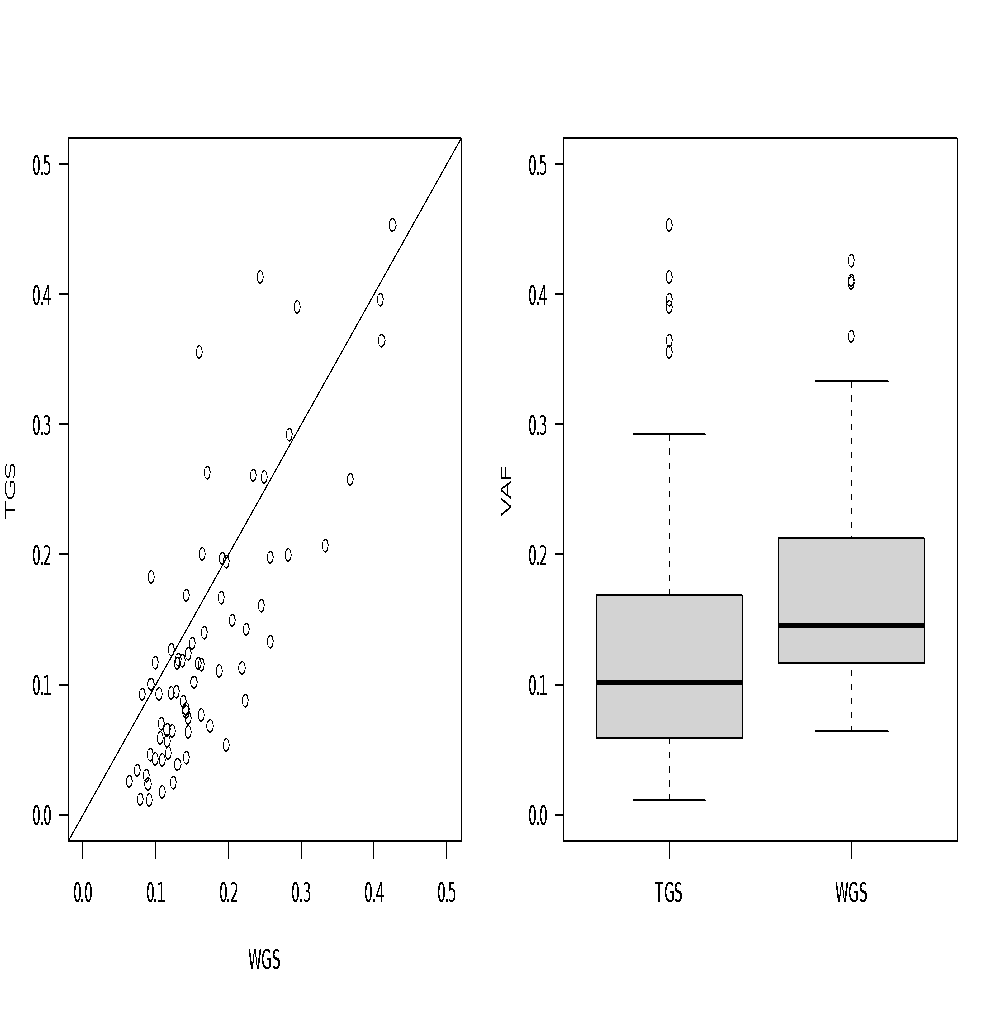

**Fig. S1.** Comparison of VAFs for CHIP mutations detected by smMIPS (y-axis) vs VAF from WGS of the same sample (x-axis); Pearson’s r=0.79.

### **Supplementary Tables**

**Table S1: Summary for 182 WHI subjects.**

|  | Overall |
| --- | --- |
| Number of individuals | 182 |
| Age at first blood draw (mean (SD)) | 64.78 (7.61) |
| BMI at first blood draw (mean (SD)) | 29.59 (5.65) |
| The first Visit (%) | |
| AV1 | 60 (33.0) |
| AV3 | 5 (2.7) |
| Base | 117 (64.3) |
| RACENIH (%) | |
| Black | 35 (19.2) |
| More than one race | 2 (1.1) |
| Unknown/Not reported | 9 (4.9) |
| White | 136 (74.7) |
| Smoking status at first blood draw (%) | |
| Current | 7 (3.8) |
| Missing | 2 (1.1) |
| Never | 107 (58.8) |
| Past | 66 (36.3) |
| Number of time points per individual (%) | |
| 1 | 8 (4.4) |
| 2 | 88 (48.4) |
| 3 | 3 (1.6) |
| 4 | 48 (26.4) |
| 5 | 27 (14.8) |
| 6 | 8 (4.4) |

**Table S2. CHIP calls in the WHI longitudinal samples.** AV: Annual visit; LLS: Long-life study visit; REF: Reference Allele; ALT: Alternative Allele; VAF: Variant allele fraction; ALT Count: number of reads supporting alternative allele.

**Table S3: Cross-sectional association at the first visit.** For model 1-4, logistic regressions are conducted on the prevalence of CHIP mutation (VAF >= 2% VS VAF <2%) against age and covariates race (Black/ White /Unknown or Other), smoking history (Past/ Never/ Current/ Missing), and BMI. For models 2 and 3, model 1 is the null model and p-values are obtained from a likelihood ratio test whether the covariate is associated with the prevalence of CHIP mutations. For model 5-8, linear regressions are conducted on the log10(VAF) of detectable CHIP mutation (VAF >0) against age and the covariate. For models 6 and 7, the model 5 is the null-model, and p-values are obtained from F tests.

| index | model | p-value |
| --- | --- | --- |
| Logistic regression (n=156) | | |
| 1 | y ~ age | 5.95e-08 |
| 2 | y ~ age + race | 0.3687 |
| 3 | y ~ age + smoking | 0.9696 |
| 4 | y ~ age + bmi | 0.174 |
| Linear regression (n=152) | | |
| 5 | y ~ age | 8.92e-08 |
| 6 | y ~ age + race | 0.207 |
| 7 | y ~ age + smoking | 0.8762 |
| 8 | y ~ age + bmi | 0.0446 |

**Table S4: Association tests of VAF changes between the first and last positive VAFs (n=148).** Linear regressions are conducted on the log10(VAF) changes of CHIP mutation against initial age, age length (age differences between the first and the last VAF samples), and covariate. For models 3 and 4, model 2 is the null model and p-values are obtained from testing whether race/smoking is associated with the prevalence of CHIP mutations. The covariates include race categories (Black/White/Unknown or other), smoking history (Never/Ever/Missing), and average BMI. For each subject carrying multiple clones, the dominant clone is selected based on the largest VAF value at any available time points.

| index | model | p-value |
| --- | --- | --- |
| 1 | y = age | 0.606 |
| 2 | y = age + age_len | 0.862 |
| 3 | y = age + age_len + race | 0.4959 |
| 4 | y = age + age_len + smoking | 0.6418 |
| 5 | y = age + age_len + bmi | 0.296 |

**Table S5**: **CHIP mutations queried in this study.** CHIP: clonal hematopoiesis of indeterminate potential; **SRSF2, IDH1, IDH2, JAK2* target a single hotspot mutation

| **Gene name** | **Reported mutations used for variant calling** | **Accession** |
| --- | --- | --- |
| ***ASXL1*** | Frameshift/nonsense/splice-site in exon 11-12 | NM_015338 |
| ***CBL*** | RING finger missense p.381-421, D348N, L380P | NM_005188 |
| ***DNMT3A*** | Frameshift/nonsense/splice-site, F290I, F290C, F290S, G293R, L295P, L295Q, L295V, V296G, V296L, V296M, W297C, W297L, W297R, W297G, G298W, G298R, G298E, W306C, P307S, P307R, P307L, P307T, G308D, I310F, I310L, I310S, I310T, S312F, R326G, R326H, R326L, R326C, R326S, V328A, V328D, V328G, F331V, G332R, G332E, S337A, S337L, S337P, V339A, V339M, V339G, L344Q, L344P, L344R, L347P, L347R, L347Q, S352N, Y365C, R366C, R366P, R366H, R366G, A368D, A368T, A368V, I369N, I369S, V372D, L373Q, A376P, A376T, A376V, R379H, R379L, R379C, R379S, D389N, I407T, I407N, I407S, W409R, A410D, A410T, G413V, F414L, F414I, F414S, F414V, F414C, A462V, K468R, E477Q, E477K, V483G, R484Q, R484W, C494Y, C497G, C497R, C497Y, G498E, H506R, G511E, C514Y, Q527H, Q527P, D529N, D529V, D531N, D531Y, Y533C, S535F, S535P, C537G, C537R, C540Y, G543A, G543S, G543C, G543D, G543V, L547H, L547P, L547F, L547R, C537Y, M548I, M548L, M548K, M548R, M548T, G542V, G550R, C554Y, R556K, R556S, R556G, C559R, C559Y, C562Y, V563M, P580L, W581R, W581G, W581C, W581S, C583S, C583Y, C586G, C586R, C586Y, K589N, L595P, R596W, R598Q, R604Q, R604W, P633H, P633L, I634F, I634T, R635G, R635L, R635P, R635W, R635Q, V636A, V636G, V636M, V636L, L637R, L637P, L637Q, S638F, S638P, S638Y, L639R, L639V, L639F, A644T, T645A, G646V, G646E, L647H, L648P, V649G, V649L, V649M, L650V, L650Q, L653W, L653F, I655N, I655T, Q656K, V657A, V657M, V657G, D658V, D658Y, R659C, R659G, R659H, Y660C, Y660N, Y660F, Y660H, Y660D, A662D, S663L, S663W, E664K, V665G, V665L, S669F, S669P, M674V, V675A, V675M, R676L, R676W, R676Q, I681N, I681S, I681M, M682R, Y683D, V684F, G685R, G685E, G685A, D686Y, D686G, D686H, D686V, V687L, D686A, V687F, R688C, R688G, R688H, V690G, V690F, V690D, T691I, I695N, H694Y, H694P, I695F, I695T, Q696P, W698C, W698R, W698S, G699R, G699S, G699D, G699V, P700L, P700S, P700R, P700Q, P700T, P700A, F701V, D702A, D702G, D702E, D702V, D702N, D702Y, L703P, L703R, L703V, V704A, V704M, V704G, I705F, I705T, I705S, I705N, G706E, G706W, G706R, G706V, G707C, G707D, G707S, G707R, G707V, C710S, C710Y, D712A, L713F, S714C, V716D, V716F, V716I, N717S, N717I, P718L, R720C, R720H, R720G, R720S, K721R, K721T, K721N, Y724C, E725K, G726V, G728D, R729Q, R729W, R729G, R729L, F731C, F731L, F731Y, F731I, F731V, F732del, F732C, F732I, F732S, F732L, F732V, E733G, E733A, E733V, F734L, F734C, F734V, Y735C, Y735N, Y735S, Y735F, Y735H, R736G, R736H, R736C, R736L, R736P, R736S, L737H, L737P, L737V, L737F, L737R, L738P, L738Q, H739P, A741G, A741V, R742L, R742G, R742P, P743H, P743R, P743L, P743S, R749C, R749L, R749H, R749G, P750R, F751L, F751C, F751I, F751V, F752del, F752C, F752L, F752I, F752V, W753G, W753C, W753L, W753R, W753S, L754P, L754R, L754H, F755S, F755I, F755L, M761I, M761V, G762C, V763G, V763I, K766E, D768E, D768H, D768V, D768Y, I769N, I769S, I769T, I769V, S770L, S770W, S770P, R771G, R771L, R771P, R771Q, F772C, F772I, F772V, L773H, L773I, L773R, L773V, E774A, E774K, E774D, E774G, E774V, S775F, S775P, P777A, P777H, P777L, P777R, P777T, P777S, V778M, I780N, I780S, I780T, D781G, V785M, A787G, A787S, H789Q, R790W, A791V, R792C, R792H, R792S, F794L, F794V, W795S, W795G, W795C, W795L, W795R, G796A, G796C, G796D, G796V, N797D, N797Y, N797H, L798P, L798H, N797K, N797S, P799L, P799A, P799T, P799S, P799R, P799H, G800S, M801I, M801R, M801T, M801V, R803S, R803G, R803K, R803T, R803W, R803M, P804L, P804S, L815Q, H821D, H821P, H821R, K826N, K826T, K826R, S828N, K829R, T835M, N838D, S839P, K841N, K841T, K841Q, Q842E, Q842R, G843S, P849L, M853R, M852L, M852V, D857N, W860R, E863D, E863G, E863K, E863V, F868S, F868L, G869S, G869V, F870V, H873R, Y874C, R879D, M880I, M880L, M880V, S881R, S881I, R882H, R882P, R882L, R882C, R882G, R882S, A884P, A884V, R885K, Q886E, Q886R, L889P, L889R, G890D, G890R, G890S, G890V, W893S, V895M, P896L, P898S, V897G, V897D, I898T, R899P, R899L, R899G, R899H, R899C, R899S, L901P, L901R, L901H, L901V, A903P, A903T, P904L, P904Q, P904A, P904R, P904S, L905R, L905P, L905Q, L905V, K906E, E907G, Y908C, Y908D, Y908N, F909C, A910P, A910V, C911R, C911Y | NM_022552 |
| ***GNB1*** | K57N, K57M, K57E, K57T, I80T, I80N | NM_002074 |
| ***IDH1**** | R132C, R132G, R132H, R132L, R132P, R132V | NM_005896 |
| ***IDH2**** | R140W, R140Q, R140L, R140G | NM_002168 |
| ***JAK2**** | V617F | NM_004972 |
| ***PPM1D*** | Frameshift/nonsense in exon 5 or exon 6 | NM_003620 |
| ***SF3B1*** | G347V, R387W, R387Q, E592K, E622D, Y623C, R625L, R625C, R625G, N626D, R630G, S637Y, H662Q, H662D, T663I, K666N, K666Q, K666T, K666E, K666R, K700E, V701F, A708T, G740R, G740E, G742D, A744P, A745P, K748E, R775P, D781G, E783K, R831Q, L833F, E862K, R957Q | NM_012433 |
| ***SRSF2**** | P95H, P95L, P95T, P95R, P95A, P95fs | NM_003016 |
| ***TET2*** | Frameshift/nonsense/splice-site, missense mutations in catalytic domains (p.1104-1481 and 1843-2002), D1121Y, D1129Y, C1133R, C1133W, C1135Y, C1135F, C1135W, G1137D, G1137V, E1137K, E1137D, E1141K, E1144K, Y1148C, L1151R, G1152R, A1153T, A1153V, G1154S, C1156Y, V1157M, I1160F, I1160S, R1161G, R1161S, M1164I, E1165K, E1165D, R1167G, R1167K, R1167S, R1167M, L1172R, A1174T, I1175T, V1180D, M1185I, E1186A, G1187S, K1188R, G1192V, C1193Y, C1193W, P1194L, P1194R, I1195V, K1197E, W1198C, W1198R, V1201I, E1207D, L1209P, L1210P, C1211Y, L1212S, V1213M, V1213E, R1214W, R1214Q, R1216Q, H1219D, H1219R, H1219Y, C1221Y, C1221R, C1221S, C1221W, C1221F, L1229R, G1235E, R1235W, L1238V, A1241S, K1243R, K1243N, L1244P, Y1245C, Y1245N, L1248P, L1248R, L1252P, L1252V, G1256C, R1261C, R1261S, R1261H, R1261P, R1262W, C1263F, C1263Y, N1266D, N1266K, N1266H, N1266Y, N1266S, C1271W, C1271S, C1273S, C1273W, C1273R, Q1274P, G1275R, G1275V, G1282R, G1282D, R1283P, S1284F, F1287V, G1288D, G1288V, C1289F, C1289W, S1290L, W1291C, S1292R, M1293I, Y1294C, G1297E, G1297R, C1298Y, C1298S, K1299M, K1299Q, K1299N, F1300V, F1300L, F1300I, S1303G, S1303R, K1310Q, L1311Q, E1318E, L1322Q, L1322P, L1322R, L1326W, L1329P, L1329Q, L1332P, M1333K, L1340R, L1340P, Y1345D, Y1345C, Q1348K, Q1348R, I1349N, E1352K, A1355V, C1358S, C1358W, R1359C, R1359L, R1359P, R1359H, R1359G, R1359S, L1360R, G1361C, G1361S, G1361D, R1366H, R1366L, R1366C, P1367L, P1367S, P1367R, F1368L, G1370V, G1370R, V1371D, A1373P, A1376V, D1376G, F1377V, F1377I, C1378R, C1378Y, C1378F, H1380Y, H1380L, H1380R, H1380Q, R1380C, H1380D, H1382R, H1382P, R1383G, H1386D, R1387H, N1387S, T1393A, C1395Y, T1397I, L1398R, F1398C, F1398L, L1398H, L1398P, H1401Y, E1401A, N1403S, Q1414R, Q1414H, Q1414K, V1416I, V1417F, P1419R, D1427V, Q1435K, V1438F, G1861E, G1861R, G1861V, A1863V, H1868Y, H1868P, H1868L, G1869W, S1870L, L1872P, I1873T, I1873S, A1876T, A1876V, R1878P, R1878H, E1879Q, E1879G, E1879D, H1881N, H1881L, T1884A, T1884I, P1889L, N1890S, P1894T, P1894R, P1894L, P1894H, R1896G, I1897N, S1898F, V1900F, E1900K, V1900D, Y1902H, Y1902C, Q1903R, A1903V, H1904R, H1904Q, K1905E, T1905A, H1912R, H1912D, H1912Y, G1913D, A1919D, F1922S, Y1923H, H1925Q, K1934N, R1966C, D1981A, S1982Y | NM_001127208 |
| ***TP53*** | Frameshift/nonsense/splice-site, S46F, G105C, G105R, G105D, G108S, G108C, R110L, R110C, T118A, T118R, T118I, S127F, S127Y, L130V, L130F, K132Q, K132E, K132W, K132R, K132M, K132N, F134V, F134L, F134S, C135W, C135S, C135F, C135G, C135Y, Q136K, Q136E, Q136P, Q136R, Q136L, Q136H, A138P, A138V, A138A, A138T, T140I, C141R, C141G, C141A, C141Y, C141S, C141F, C141W, V143M, V143A, V143E, L145Q, W146C, W146L, L145R, V147G, P151T, P151A, P151S, P151H, P151R, P152S, P152R, P152L, T155P, T155A, V157F, R158H, R158L, A159V, A159P, A159S, A159D, A161T, A161D, Y163N, Y163H, Y163D, Y163S, Y163C, K164E, K164M, K164N, K164P, H168Y, H168P, H168R, H168L, H168Q, M169I, M169T, M169V, E171K, E171Q, E171G, E171A, E171V, E171D, V172D, V173M, V173L, V173G, R174W, R175G, R175C, R175H, C176R, C176G, C176Y, C176F, C176S, P177R, P177L, H178D, H178P, H178Q, H179Y, H179R, H179D, H179Q, R181C, R181Y, R181H, D186G, G187S, P190L, P190T, H193N, H193P, H193L, H193R, L194F, L194R, I195F, I195N, I195T, R196P, V197L, G199V, Y205D, Y205N, Y205C, V203M, Y205H, D208V, R213Q, R213P, F212I, R213L, R213Q, H214D, H214P, H214R, S215G, S215I, S215R, V216M, V217G, Y220N, Y220H, Y220S, Y220C, E224D, I232F, I232N, I232T, I232S, Y234N, Y234H, Y234S, Y234C, Y236N, Y236H, Y236C, M237V, M237K, M237I, C238R, C238G, C238Y, C238W, N239T, N239S, S241Y, S241C, S241F, C242G, C242Y, C242S, C242F, G244S, G244C, G244D, G245S, G245R, G245C, G245D, G245A, G245V, G245S, M246V, M246K, M246R, M246I, N247I, R248W, R248G, R248Q, R249G, R249W, R249T, R249M, P250L, I251N, L252P, I254S, I255F, I255N, I255S, L257Q, L257P, E258K, E258Q, D259Y, S261T, G262D, G262V, L265P, G266R, G266E, G266V, R267W, R267Q, R267P, E271K, V272M, V272L, R273S, R273G, R273C, R273H, R273P, R273L, V274F, V274D, V274A, V274G, V274L, C275Y, C275S, C275F, A276P, C277F, C277Y, P278T, P278A, P278S, P278H, P278R, P278L, G279E, R280G, R280K, R280T, R280I, R280S, D281N, D281H, D281Y, D281G, D281E, D281V, R282G, R282W, R282Q, R282P, E285K, E285V, E286G, E286V, E286K, K320N, L330R, G334V, R337C, R337L, A347T, L348F, T377P | NM_001126112 |
| ***U2AF1*** | D14G, S34F, S34Y, R35L, R156H, R156Q, Q157R, Q157P | NM_006758 |
| ***ZBTB33*** | Frameshift/nonsense/splice-site, A9P, D11V, D11G, S15C, L19P, Q25E, Q25R, R26C, R26H, C32G, C32Y, V34G, V34D, T35P, R41Q, A45P, L50I, S54G, Y56C, Q59P, Q59K, V67I, V68D, L70R, R74G, I77F, I77N, E80D, Y86C, I90N, E100G, L108V, I113K, I113T, A114T, A114V, L116H, Y434H, Y434C, A435V, I437T, G438S, G438V, T441A, Y442C, D443G, I446S, P447A, L459F, E485D, Y494C, C496R, C496F, C496S, C496Y, R501T, Y503S, L509V, H512Y, H512R, N514K, S517C, Y522C, Y522H, R525C, Y526N, Y526D, C527R, C527Y, P532S, L533F, A534V, A534T, E535Q, E535K, E535A, R537C, R537H, R537L, T538A, T538R, H540N, E541K, H543D, H544R, E547K, R549K, Y550C, Q551P, C552G, C552Y, C555Y, Y562C, Q563P, S566P, Y584C, S591A | NM_006777 |
| ***ZNF318*** | Frameshift/nonsense/splice-site | NM_014345 |
